# Parkinson Disease Neuropathological Comorbidities: Prevalences from Younger-Old to Older-Old, With Comparison to Non-Demented, Non-Parkinsonian Subjects

**DOI:** 10.1101/2025.01.13.25319971

**Authors:** Thomas G. Beach, Geidy E. Serrano, Erika D. Driver-Dunckley, Lucia I. Sue, Holly A. Shill, Shyamal H. Mehta, Christine M. Belden, Ileana Lorenzini, Cecilia Tremblay, Parichita Choudhury, Alireza Atri, Charles H. Adler

## Abstract

Co-existing neuropathological comorbidities have been repeatedly reported to be extremely common in subjects dying with dementia due to Alzheimer disease. As these are likely to be additive to cognitive impairment, and may not be affected by molecularly-specific AD therapeutics, they may cause significant inter-individual response heterogeneity amongst subjects in AD clinical trials. Furthermore, while originally noted for the oldest old, recent reports have now documented high neuropathological comorbidity prevalences in younger old AD subjects, who are more likely to be included in clinical trials. Comorbid neuropathologies in subjects with Parkinson disease have received much less attention. As with AD, comorbidities may interfere with the evaluation of PD clinical trials. We have here examined the decadal-wise presence of multiple co-pathologies, and their clinical effects, in a series of autopsies of PD and control subjects from the Arizona Study of Aging and Neurodegenerative Disorders. Amyloid plaques were present in more than 40% of PD patients in their 60s, and in 85% of those in their 90s. Neurofibrillary tangles were present in PD, as in all elderly humans, from the 60s onwards, while Braak tangle stages of IV or greater, which are strongly associated with severe cognitive impairment, reached 40%, 50% and 60% in those subjects in their 70s, 80s and 90s, respectively. Both plaques and tangles were significant predictors of a lower MMSE score while greater CAA scores had borderline significance. None of these, however, were independently associated with a higher UPDRS score. The ApoE4 allele, while a known predictor of AD pathology, especially amyloid plaques, was not an independent predictor of function with either of these clinical measures, suggesting that its influence is largely mediated by the neuropathologies that it predisposes to. Non-AD tauopathies, including the major conditions of progressive supranuclear palsy (PSP), and corticobasal degeneration (CBD), as well as the microscopic changes of argyrophilic grains (ARG) and aging-related tau astrogliopathy (ARTAG), also co-existed with PD, especially ARG and ARTAG, which ranged between 20% and 40% across decades for the former and up to 80% for the latter; we did not find significant associations of either with final MMSE or UPDRS scores. We failed to find a significant association of limbic TDP-43 histopathology with either final MMSE score or final UPDRS motor score but our subjects were more heavily weighted with PD than those in prior studies and our cognitive correlate, limited to MMSE score, may have missed associations with cognitive domain subsets. We found several cerebrovascular pathologies to be predictors of both cognitive and motor impairment, including brain infarcts, circle of Willis atherosclerosis, and higher white matter rarefaction score. All of the investigated pathology types were common in the non-demented, non-parkinsonian control subjects, and all increased with age in parallel with those co-existing with PD, while with generally lower prevalences. The high concurrence rate of the neurodegenerative protein aggregate diseases is suggestive of either a synergistic co-pathogenesis, where one aggregate type may instigate or accelerate another type, or of one or more underlying predisposing physiological or molecular mechanisms.

## Introduction

Co-existing neuropathological comorbidities, including insoluble aggregates of Aβ, tau, α-synuclein, and TDP-43, as well as a variety of cerebrovascular lesions, are ubiquitous in the brains of elderly subjects with the major neurodegenerative diseases. This has been most comprehensively documented for subjects whose primary pathology is Alzheimer’s disease (AD), in whom they are likely to at least be additive to the rate of cognitive decline or to the ultimate severity of cognitive impairment [1–10]. While originally noted for the “oldest old” [10], recent reports document high neuropathological comorbidity prevalences in “younger old” AD subjects as well, including early-onset sporadic and autosomal dominant inherited AD [11–13], raising potential difficulties for clinical trials, which selectively target these younger age groups.

The panoply of comorbid neuropathologies present in subjects with Parkinson’s disease (PD), as well as their impact on both motor and cognitive function, has received much less attention [14–18], as has their presence in unimpaired elderly individuals [19–21]. Comorbidities may interfere with the evaluation of PD clinical trials as they may not respond to α-synuclein-targeted molecular therapeutics, and their presence in clinically normal elderly people suggests that they are likely to be present even before typical signs and symptoms emerge. At present, PD clinical trials are only able to stratify for AD presence as there are no proven sensitive and specific *in vivo* diagnostics for the many other pathology types that might be co-existing. It is therefore critical that we assess the probabilities of their age-related co-existence in persons with PD. We here examine the decadal-wise presence of multiple co-pathologies in a series of PD autopsies from the Arizona Study of Aging and Neurodegenerative Disorders (AZSAND)[5], a longitudinal clinicopathological study with a focus on both AD and PD as well as normal aging.

## Materials and Methods

### Subject selection

Parkinson’s disease (PD) and control subjects for comorbidity prevalence determination were selected by database searches of the AZSAND/Brain and Body Donation Program (www.brainandbodydonationprogram.org) [5]. Search criteria specified that subjects died and had a complete neuropathological examination with a clinicopathological diagnosis of PD or PD with dementia, or were non-demented, non-parkinsonian controls without Lewy pathology. Diagnostic criteria for PD included having had two of three cardinal signs (bradykinesia, rest tremor, cogwheel rigidity) of parkinsonism as well as substantia nigra (SN) pigmented neuron loss and Lewy-type synucleinopathy (LTS) in the SN at autopsy [22]. Subjects classified as AD had dementia and met Intermediate or High National Institute on Aging-Reagan Institute (NIA-RI) and/or NIA-Alzheimer’s Association AD neuropathological criteria [23,24]. Control subjects were defined as those that were not clinically diagnosed with dementia or parkinsonism during life, had no LTS and did not meet clinicopathological diagnostic criteria for any defined major neurodegenerative condition. All subjects were subdivided by age at death into those that died in their 50s/ 60s, 70s, 80s or 90s/100s.

Subjects for logistic regression analysis of the effects of neuropathological lesion type on cognitive and motor function were also selected by AZSAND database searches. To increase subject numbers and statistical power for TDP-43 pathology, selection criteria were opened to included diagnoses of all types, provided that they had an apoliprotein E (ApoE) genotype determination, a final Mini Mental State Examination (MMSE) score, a final Unified Parkinson’s Disease Rating Scale (UPDRS) motor score, and TDP-43 neuropathological assessment.

### Subject characterization

All subjects had at least one, and in most cases annual, standardized research-dedicated AZSAND clinical evaluations, done by teams of nurses, medical assistants, behavioral neurologists, movement disorders neurologists, neuropsychologists and psychometrists using standardized assessment batteries [5], including but not restricted to, the MMSE, National Alzheimer’s Coordinating Center (NACC) Uniform Data Set (UDS) and the UPDRS.

All subjects received identical blinded neuropathological examinations [5] by a single observer (TGB), including summary regional brain density measures for total amyloid plaques and neurofibrillary tangles (for both is a summary score of 5 regional semi-quantitative 0-3 density scores for a maximum possible total of 15 in frontal, temporal and parietal lobes plus hippocampal CA1 and entorhinal/transentorhinal area), summary LTS regional brain density scores (summary score of semi-quantitative 0-4 density scores in 10 brain regions for a maximum possible total of 40), and staging using the Unified Staging System for Lewy Body Disorders [25], as well as substantia nigra depigmentation scores (0-3 for none, mild, moderate and severe) and assignment of CERAD neuritic plaque density, Braak neurofibrillary stage, and AD neuropathological change levels of Low, Intermediate or High, as described previously [23,24]. Cerebrovascular-related neuropathological variables included total cerebral amyloid angiopathy (CAA) score and total cerebral white matter rarefaction (WMR) score (for both is the summary score of 4 regional semi-quantitative 0-3 density scores for a maximum possible total of 12 in frontal, temporal, parietal and occipital lobes), as well as circle of Willis atherosclerosis score (0-3 for none, mild, moderate and severe) and presence or absence of any brain infarcts (including both macroscopic and microscopic brain infarcts). Pathological TDP-43 deposits were immunohistochemically detected and semi-quantitatively assessed (0-3 for none, sparse, moderate and severe), in sections of amygdala, hippocampal CA1, entorhinal/transentorhinal area, middle temporal gyrus and middle frontal gyrus, with antibodies to TDP-43 phosphorylated at phosphoserine residues 409-410 as previously described [26,27]. The distinction between LATE (limbic associated TDP-43 proteinopathy) and FTLD-TDP (frontotemporal lobar degeneration with TDP-43 proteinopathy) was made pragmatically, on the basis of the presence or absence of phosphorylated TDP-43 pathology in frontal and temporal neocortex (middle frontal gyrus and middle temporal gyrus).

### Statistical analysis

To determine the statistical significance of comorbidity prevalences, proportions of subjects across decades (50s/60s, 70s, 80s and 90s/100s) and diagnoses (PD vs Control) with each pathology type were compared using Chi-square tests or Fisher’s Exact tests as appropriate. The objectives were to determine whether age groups and/or PD and control cases were significantly different in terms of the presence of comorbidity types.

We also used logistic regression models adjusted for age, sex, and possession of an apolipoprotein E-ε4 allele (ApoE4), to assess the independent influence of the specific neuropathological lesion types on cognitive and motor impairment, as documented by the final Mini Mental State Examination (MMSE) and UPDRS motor scores, respectively.

## Results

### Subjects Selected for Study

Database queries resulted in 269 PD subjects and 232 Control subjects (Supplementary Tables 1 and 2, respectively) for the comparison of comorbidity prevalences across decades, and 576 subjects (Supplementary Table 3) for the logistic regression analyses of comorbidity type association with MMSE and UPDRS motor scores. Due to the later world-wide recognition of the existence of TDP-43 and ARTAG pathology types, ARTAG data for comparison of comorbidity prevalences was missing for 178 and 154 PD and control cases, while TDP-43 data was missing for 152 and 110 PD and Control cases, respectively. Because of this and absence of its effect on statistical power in preliminary models, ARTAG was not included as a variable in the logistic regression analyses while all 576 cases had TDP-43 data available. The dataset for logistic regression analysis included, along with 88 PD and 204 Control subjects, 225 subjects neuropathologically diagnosed with AD, 47 with vascular dementia, 31 with dementia with Lewy bodies, 18 with incidental Lewy body disease, 31 with progressive supranuclear palsy, 7 with multiple system atrophy, and 11 with FTLD-TDP; multiple diagnoses were extremely common (see Supplementary Table 3 for complete lists of diagnoses for each case). For all Supplementary Tables, the exact age at death is not given due to privacy concerns. Case IDs and Donor IDs are known only to researchers of AZSAND/BBDP; they are not accessible to hospital or healthcare staff.

### PD Comorbidity Prevalences by Decade of Death, in Comparison with Control Cases

Figure 1 A-D shows graphs depicting percentages of neuropathological comorbidity types in clinicopathologically-diagnosed PD cases, by decade of death age, with Chi-square or Fisher’s Exact Test analysis of differences between decades. Almost all comorbidities increased in prevalence over successive decades. For AD-related comorbidity types (A), these ranged from roughly 40% to 80% for amyloid plaques, 20% to near 80% for CAA, and from 6% to near 60% for Braak tau/tangle stage of IV or greater. Of non-AD tauopathies (B), ARTAG prevalences ranged from about 20% to 80%, while argyrophilic grains (ARG) ranged from 12% to 45%. Progressive supranuclear palsy (PSP) microscopic changes were found in 6% of the youngest subjects and 15% in the oldest group. Only two subjects in total had microscopic changes of corticobasal degeneration (CBD); none had changes of Pick’s disease. Of TDP-43 proteinopathies and related conditions (C), including LATE, hippocampal sclerosis, FTLD-TDP and motor neuron disease, only LATE was common, ranging from around 20% in the youngest cases to 50% in the oldest group. Hippocampal sclerosis was present in only 8 PD cases, most of which were in the oldest age group; TDP-43 pathology was present in 7 of these cases. No PD cases had co-existing FTLD-TDP or motor neuron disease. Cerebrovascular disease type prevalences (D) commonly seen in PD cases included notable circle of Willis atherosclerotic stenosis (grade of moderate or severe) and at least mild to moderate cerebral white matter rarefaction (WMR), both present from about 30% in the youngest category to 70-75% in the oldest. Cases with any number of microscopic infarcts ranged from 25% to 50% while those with any number of macroscopic infarcts stayed relatively constant across the decades, between 10% and 20%.

**Figure 1.**
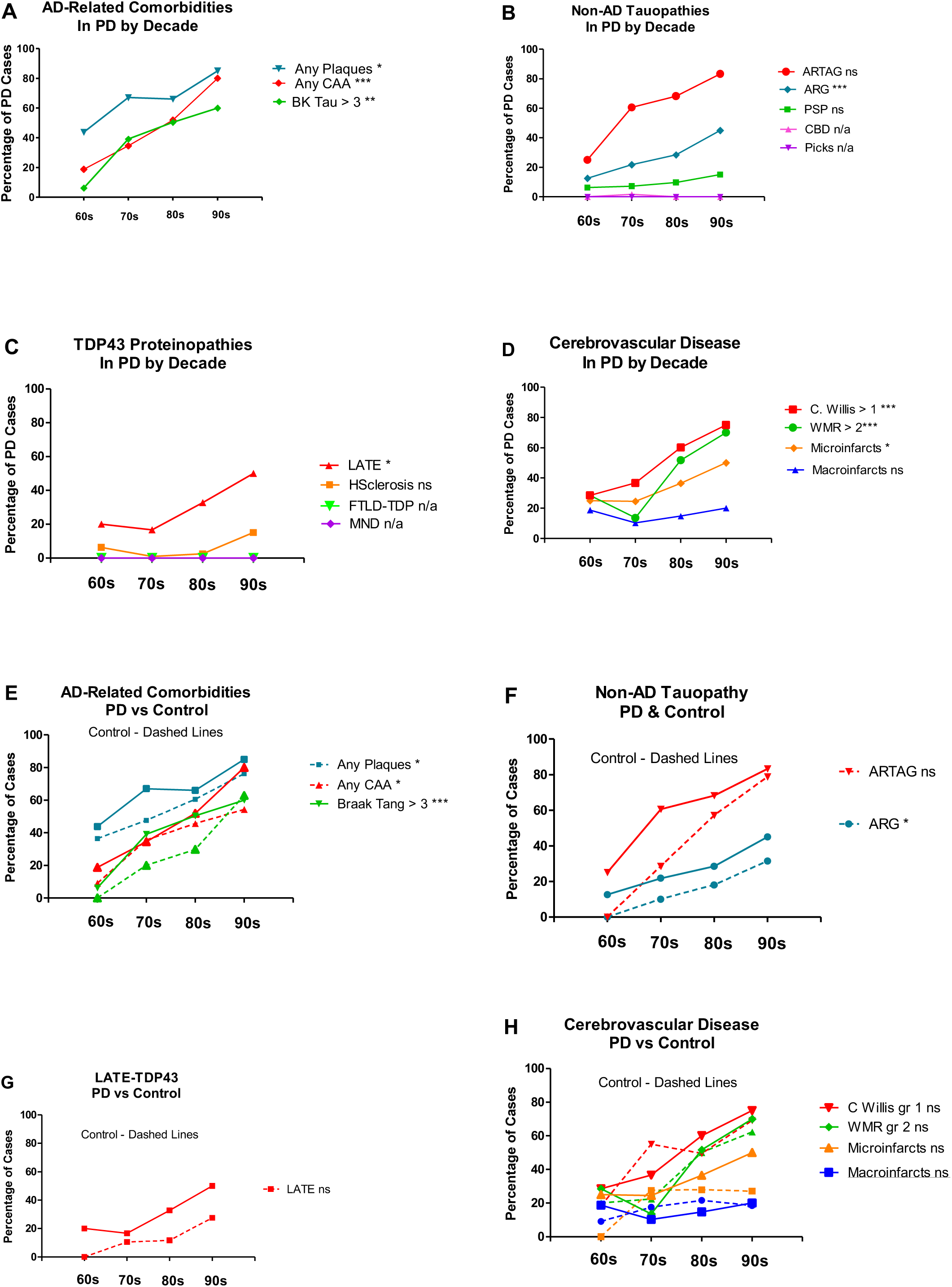
Figure 1 A-D shows graphs depicting percentages of neuropathological comorbidity types in clinicopathologically-diagnosed PD cases, by decade of death age, with Chi-square or Fisher’s Exact Test analysis of differences between decades. Figure 1 E-H show duplicate graphs from A through D with added dashed lines for percentages of the same PD neuropathological comorbidity types that are also present in control subjects. Chi-square or Fisher’s Exact Tests were done to determine significant differences between PD and control cases. For all comparisons: * = p < 0.05; ** = p < 0.01; *** = p < 0.001.

Figure 1 E-H show duplicate graphs from A through D with added dashed lines for percentages of the same PD neuropathological comorbidity types that are also present in control subjects. All comorbidities except cerebrovascular types (H) showed a general trend for lesser prevalences in control cases. Significant proportional differences between PD and control cases are present for plaques in subjects in their 70s, for CAA in subjects in their 90s, for Braak tau/tangle stage greater than 3 in subjects in their 80s and for ARG when data is combined across age groups.

### Cognitive and Motor Effects of Comorbidities

Logistic regression analysis was used to determine the independence and significance of neuropathology types on motor and cognitive function as reflected by the final MMSE and UPDRS motor scores. To simplify comparison of odds ratios (ORs), all variables, except for age, sex, and interval between last MMSE or UPDRS score, were expressed as binary measures based on decisions as indicated in Table 1. As preliminary models indicated a lack of significant associations with PSP, argyrophilic grains or ARTAG, these were not included in the final analyses.

**Table 1.** Variables for logistic regression analyses of the independent effects of neuropathological lesion types on cognitive and motor function status. To simplify comparison of odds ratios, all variables, except for age and interval between last MMSE or UPDRS score, were expressed as binary measures based on decisions as indicated. Male sex = 1.

| Variable | Abbreviation | Cut Point | Explanation/Rationale |
| --- | --- | --- | --- |
| Last MMSE Score | MMSE | < 24 = 1 | Roughly consistent with dementia |
| Last UPDRS Score | UPDRS | > 13 = 1 | 13 is median UPDRS motor score for all cases |
| Apolipoprotein E4 | APOE4 | Any E4 = 1 | Prior literature |
| Total Plaque Score | PLQgr5 | > 5 = 1 | Score of 6/15 or more indicates an average plaque density of at least moderate across frontal, parietal and temporal neocortex |
| Total Tangle Score | TANGgr5 | > 5 = 1 | Score of 6/15 or more indicates an average neurofibrillary tangle density score of at least moderate across hippocampus and entorhinal/transentorhinal areas |
| Total CAA Score | CAAgr4 | > 4 = 1 | Score greater than 4/12 indicates an average CAA score of at least sparse to moderate across frontal, parietal, temporal and occipital neocortex |
| LB Synucleinopathy | LBgr12 | > 12 = 1 | Score greater than 12/40 indicates an average synucleinopathy density score of at least sparse to moderate across olfactory bulb, medulla, pons and amygdala |
| SN Depigmentation | SNdepg3 | Score 3 = 1 | Score of 3/3 indicates severe loss of SN pigmented neurons |
| C. Willis Atherosclerosis | CW3 | Score 3 = 1 | Score of 3/3 indicates severe Circle of Willis atherosclerosis |
| White Matter Rarefaction | WMgr4 | > 4 = 1 | Score greater than 4/12 indicates an average score of at least mild to moderate across frontal, parietal, temporal and occipital neocortex |
| TDP-43 Proteinopathy | TDP43 | Any = 1 | Any TDP-43 proteinopathy across amygdala, hippocampal formation or parahippocampal gyrus |
| Any Brain Infarct | InfANY | Any = 1 | Any macroscopic or microscopic infarct in any brain region |

For a higher final UPDRS motor score, significant predictive variables included shorter test-to-death interval, higher LTS total brain score, higher white matter rarefaction (WMR) score, and any brain infarct (Table 2). Of these, a higher LTS total brain score had the strongest and most significant predictive value, with an OR of 4.0. A severe circle of Willis atherosclerosis score had borderline significance, with an OR of 1.5 and p-value of 0.058. A “severe” SN pigmented neuron loss score was not a significant predictor of higher UPRDS score in the main model but was significant in a separate model (results not shown) when expressed as the full-scale score of 0-3, with an OR of 1.64; in this model, however, the LTS total brain score then failed to meet the significance level, possibly due to co-linearity.

**Table 2.** Multivariable logistic regression models assessing predictors of a higher final UPDRS motor score or lower final MMSE score. For sex, Male = 1. Interval is the interval in months between the final clinical test and death (see Supplementary <u>Data</u> File 3 for all data). For a higher final UPDRS motor score, significant predictive variables included shorter test-to-death interval, higher Lewy-type synucleinopathy total score, higher white matter rarefaction score, and any brain infarct. For a lower final MMSE score, significant predictive variables included shorter test-to-death interval, higher plaque and tangle density scores, higher Lewy-type synucleinopathy total score, higher Circle of Willis atherosclerosis score, and higher white matter rarefaction score. * p < 0.05; ** p < 0.01; *** p < 0.001

| Variable | UPDRS Motor Score > 13 |  | MMSE Score < 24 |  |
| --- | --- | --- | --- | --- |
|  | OR (95% CI) | P value | OR (95% CI) | P value |
| Interval | 0.9740 (0.963, 0.986) | 0.000012*** | 0.9826 (0.973, 0.992) | 0.000469*** |
| Age | 0.9560 (0.929, 0.983) | 0.001740** | 0.9283 (0.901, 0.957) | 0.000001*** |
| Sex | 1.0650 (0.793, 1.431) | 0.6760 | 1.029 (0.92, 1.151) | 0.6194 |
| APOE4 | 0.9999 (1, 1) | 0.05541 | 1.0000 (1, 1) | 0.1824 |
| PLQgr5 | 1.027 (0.679, 1.551) | 0.9011 | 1.692 (1.101, 2.599) | 0.016335* |
| TANGgr5 | 1.304 (0.853, 1.995) | 0.2201 | 3.365 (2.112, 5.36) | < 0.000001*** |
| CAAgr4 | 1.039 (0.656, 1.647) | 0.8698 | 1.522 (0.956, 2.422) | 0.07676 |
| LBgr12 | 4.005 (2.615, 6.133) | < 0.000001*** | 1.857 (1.218, 2.829) | 0.003983** |
| SNdepg3 | 1.002 (0.982, 1.023) | 0.8385 | 1.006 (0.983, 1.028) | 0.6225 |
| CW3 | 1.508 (0.986, 2.305) | 0.0580 | 2.050 (1.319, 3.186) | 0.001425** |
| WMgr4 | 1.777 (1.187, 2.661) | 0.00524** | 2.691 (1.769, 4.093) | 0.000004*** |
| TDP43 | 0.9134 (0.598, 1.394) | 0.6748 | 1.126 (0.723, 1.754) | 0.5991 |
| InfANY | 1.626 (1.073, 2.466) | 0.0220* | 1.126 (0.957, 2.256) | 0.5991 |

For a lower final MMSE score, significant predictive variables included shorter test-to-death interval, higher plaque and tangle density scores, higher LTS total brain score, higher circle of Willis atherosclerosis score, and higher WMR score (Table 2). Of these, a higher neurofibrillary tangle density total brain score had the strongest and most significant predictive value, with an OR of 3.36; higher WMR score and higher circle of Willis atherosclerosis scores followed this, with ORs of 2.6 and 2.0, respectively. Again, LTS total brain score was a significant predictor, with an OR of 1.8 but a “severe” substantia nigra pigmented neuron loss score was not a significant predictor when included in the same model. In a separate model (results not shown), the full-scale SN pigmented neuron loss score was a significant predictor, with an OR of 1.24; in this model, however, the LTS total brain score again failed to meet the significance level.

As TDP-43 pathology, when utilized as a simple present or absent binary variable, was not a significant predictor of either MMSE or UPDRS final scores, we used semi-quantitative TDP-43 density data to further test its clinical significance. For these models, a separate file (see second sheet in Supplementary Table 3) was created with subject data (n = 452) restricted to those that had TDP-43 pathology density scores for amygdala but did not have a diagnosis of FTLD-TDP and did not have TDP-43 pathology in the frontal neocortex. The semi-quantitative TDP-43 amygdala density scores were separately used as full-scale (0-3) or binary variables (greater than score 1 and greater than score 2). In none of these additional models did TDP-43 pathology reach independent significance as a predictor of either lower MMSE or higher UPDRS scores. The highest OR reached for predicting a lower MMSE score, with the binary positive score set at 3/3, was 1.55 (95% CI of 0.874 to 2.75; p = 0.133), while for the prediction of a higher UPDRS score, the highest OR was lower, at 1.190, also reached with the binary positive score set at 3/3 (95% CI of 0.685 to 2.066; p = 0.65).

## Discussion

Co-existing neuropathological comorbidities have been repeatedly reported to be so common as to be expected in the great majority of subjects dying with dementia due to Alzheimer’s disease. As these are likely to be additive to cognitive impairment [1–10], and may not be affected by molecularly-specific AD therapeutics, they may cause significant inter-individual response heterogeneity amongst subjects in AD clinical trials. Furthermore, while originally noted for the “oldest old” [10], recent reports [11–13] have now documented high neuropathological comorbidity prevalences in “younger old” AD subjects, who are more likely to be included in clinical trials.

Comorbid neuropathologies in subjects with Parkinson’s disease, as well as their age distribution and impact on both motor and cognitive function, have received much less attention [14–18], as has their presence in unimpaired elderly individuals [19–21]. As with AD clinical trials, comorbidities may interfere with the evaluation of PD clinical trials as they may interfere with response to molecularly-specific therapeutics. Furthermore, as shown in this study, they are also common in clinically normal elderly people, suggesting that they are likely to be present even before typical signs and symptoms emerge. Currently, of these comorbidities, it is only possible to stratify PD trials for AD co-existence due to the lack of proven sensitive and specific diagnostics for the multiple other pathology types. It is therefore critical that we assess, across a wide range of human aging, the probabilities that these comorbidities exist in persons with PD. We have here examined the decadal-wise presence of multiple co-pathologies, and their clinical effects, in a series of autopsies of PD and control subjects from the Arizona Study of Aging and Neurodegenerative Disorders [5].

As we and others have reported [1–4,11–21, 28], the co-existence of AD and LTS is extremely common and is likely to be clinically significant, even in subjects in their 50s and 60s and including in those early-onset genetic forms of AD including presenilin-1 mutations and Down’s syndrome. In the current study, amyloid plaques were present in more than 40% of PD patients in their 60s, and in 85% of those in their 90s. Neurofibrillary tangles were present in PD, as in all elderly humans, from the 60s onwards, while Braak tangle stages of IV or greater, which are strongly associated with severe cognitive impairment, reached 40%, 50% and 60% in those subjects in their 70s, 80s and 90s, respectively. Both plaques and tangles were significant predictors of a lower MMSE score while greater CAA scores had borderline significance. None of these, however, were independently associated with a higher UPDRS score, largely in agreement with a previous report [16], although differing in that this previous report did find CAA to be a significant predictor of progressive parkinsonism. The ApoE4 allele, while a known predictor of AD pathology, especially amyloid plaques, was not an independent predictor of function with either of these clinical measures, suggesting that its influence is largely mediated by the neuropathologies that it predisposes to, although others utilizing larger datasets have supported its independence, at least with respect to cognitive function [29].

Not as often investigated in either PD or AD has been the co-existence of non-AD tauopathies, including the major conditions of progressive supranuclear palsy (PSP), corticobasal degeneration (CBD), and Pick’s disease, as well as the microscopic changes of argyrophilic grains (ARG) and aging-related tau astrogliopathy (ARTAG). Here, similarly to what we reported for AD [11], we found a relatively low concurrence of PSP with PD, with only 5-10% overlap until ages 90 and over, when co-existence reaches 15%. We found only two subjects with neuropathological changes of both PD and CBD, and no PD subjects with Pick’s disease. In contrast, ARG and ARTAG were much more common in PD, ranging between 20% and 40% across decades for the former and up to 80% for the latter; in common with other reports [14,30,31], we did not find significant associations of either with final MMSE or UPDRS scores, although our results for ARTAG may have been affected by the relatively fewer numbers of subjects that had been assessed for it. Recent studies reported significant ARTAG associations with faster cognitive decline or greater cognitive impairment in subjects with frontal cortex ARTAG [32] and primary age-related tauopathy [33], respectively. Similarly, although most studies have reported non-significant associations of ARG with cognition, there may be a significant relationship in subjects with low to moderate Braak tangle stages [34].

The presence and clinical significance of LATE in PD has not often been previously assessed in large datasets. While LATE has repeatedly been found to be a significant independent associate of cognitive impairment and/or decline in older subjects [21,31, 35], there appear to be only two studies, both negative, assessing its relationship with parkinsonism [16,36]. We failed to find a significant association of limbic TDP-43 histopathology with either final MMSE score or final UPDRS motor score but our subjects were more heavily weighted with PD than those in prior studies and our cognitive correlate, limited to MMSE score, may have missed associations with cognitive domain subsets.

We found several cerebrovascular pathologies to be significant predictors of both cognitive and motor impairment. As for the other pathology types that we assessed, cerebrovascular disease has been extensively reported to have a significant contribution to cognitive impairment and decline rate, as recently reported in large datasets [7,21], but there are fewer extensive studies of its role in PD subjects. One study of more than 1,000 decedents found macroinfarcts, large-vessel disease (atherosclerosis) and small-vessel disease (arteriolosclerosis) all had significant contributions to a progressive parkinsonism syndrome [16]. Supporting this finding, we found that any brain infarct (combining those due to large- and small-vessel disease) and a higher WMR score (associated with small-vessel disease) both had significant predictive value for a higher UPRDS motor score, while the circle of Willis atherosclerosis score, a measure of large-vessel disease, had borderline significance (p = 0.058). In agreement with many previous studies, our indicators of both large- and small-vessel disease processes were significant predictors of a lower cognitive status.

All of the investigated pathology types were common in the non-demented, non-parkinsonian control subjects, and all increased with age in parallel with those co-existing with PD. For all pathologies and age groups except the cerebrovascular ones, the proportions of those affected were always less in the control group as compared to the PD subjects, suggesting, as others have proposed [20], that there is a synergistic relationship between protein aggregate diseases. The relationship of cerebrovascular disease with neurodegenerative disease is more complex, however, and may change over the decades from middle age to old age. Thus, while cerebrovascular disease and risk factors in middle age appear to increase risk for cognitive impairment in old age, once cognitive impairment becomes evident, or even prior to that, this relationship often becomes reversed, possibly due to weight loss brought on by AD pathology affecting the hypothalamic appetite centers [37,38]. For PD, we and others [39,40] have reported overall decreased associations with cardiovascular risk factors and their manifest expressions, which we have hypothesized may be due to lower rates of hypertension resulting from an effective ganglionic synucleinopathy-associated sympathectomy [39].

The high concurrence rate of the neurodegenerative protein aggregate diseases is suggestive of either a synergistic co-pathogenesis, where one aggregate type may instigate or accelerate another type, or of one or more underlying predisposing physiological or molecular mechanisms. We and others have postulated that both tauopathies and synucleinopathies may be to some extent secondary to cerebral Aβ plaque deposition [13,28]. In fact, both tau and synuclein pathology clearly occur as responses to several different inherited cerebral amyloidoses, including genetic early-onset forms depositing not only Aβ (PSEN1 and APP mutations, trisomy 21) but also prion protein amyloid (in Gerstmann-Straussler-Scheinker disease) [13]. The amygdala, in particular, may commonly be affected by all four major aggregate types and single neurons have been shown to harbor both Lewy bodies and neurofibrillary tangles, and both phosphorylated TDP-43 and phosphorylated tau inclusions [20,31,41–43].

Possible common underlying molecular mechanisms must include aging, most probably affecting numerous separate pathways [31]. More specific processes might involve the apoE4 allele [44] and other genetic variants, especially those affecting autophagy and lysosomal function [45,46]. Relatively neglected has been the effect of numerous implicated environmental exposures, especially for predominantly non-genetically-determined conditions including Parkinson’s disease [47].

## Data Availability

All data produced in the present work are contained in the manuscript and supplemental files.

